# Correlation of the global spread of coronavirus disease-19 with atmospheric air temperature

**DOI:** 10.1101/2020.05.27.20115048

**Authors:** Yihienew M. Bezabih, Alemitu Mequanint, Endalkachew Alamneh, Alemayehu Bezabih, Wilber Sabiiti, Anna Roujeinikova, Woldesellassie M. Bezabhe

**Author notes:** Corresponding author: Yihienew Bezabih.

## Abstract

Severe acute respiratory syndrome coronavirus 2 (SARS-CoV-2) is an enveloped virus that may be sensitive to heat. We assessed whether the spread of coronavirus disease 2019 (COVID-19) correlates with air temperature. We also studied whether additional climate, geographical, and population variables were correlated. The total number of confirmed COVID-19 cases and mortality rates reported in each country between 1^st^ Jan and 31^st^ Mar 2020 were compared with the country’s three-month average atmospheric air temperature, precipitation and latitude. Spearman’s correlation coefficient (ρ) was used to identify significant correlations. Our analysis included a total of 748,555 confirmed COVID-19 cases worldwide. The total number of patients with COVID-19 decreased with increasing atmospheric air temperature (ρ = –0.54, 95%CI: [-0.64, –0.42]; P <0.001) and increased with an increasing latitude (ρ = 0.60, 95%CI: [0.48, 0.70]; P <0.001). Our findings justify further studies to examine the effect of air temperature on infectivity of SAR-CoV-2.

## 1. Introduction

Severe acute respiratory syndrome coronavirus 2 (SARS-CoV-2) is an RNA virus that has caused the current coronavirus disease (COVID-19) pandemic **[1]**. First recorded in late 2019 in Wuhan (China), SARS-CoV-2 infection has spread to most countries around the world **[2]**. As of 31^st^ March 2020, there were 748,555 confirmed COVID-19 cases with 50,325 deaths(5.4% mortality rate) worldwide **[2]**. As a respiratory pathogen, the virus has been able to reach every corner of the world but shows strikingly different rates of spread in different countries**[2]**. Country-to-country variation in the rate of SARS-CoV-2 spread may be related to variations in climate, as the structure of the viral particles is not stable and may be sensitive to heat and/or humidity of the environment outside a host **[3]**.

Structurally, SARS-CoV-2 is composed of an outer envelope studded with projecting glycoproteins and a helical nucleocapsid at its core **[4,5]**. The nucleocapsid consists of the genomic RNA and a nucleocapsid protein (N), whereas the envelope contains E (envelope), M (membrane) and S (spike) proteins **[5]**. The S protein of the virus attaches to the angiotensin converting enzyme receptors for entry into the cells of the human respiratory tract **[6,7]**. As seen in other viruses, the envelope structures of SARS-CoV-2 are sensitive to physical and chemical conditions and can be destabilised or damaged by heat, ultraviolet (UV) light, or extreme pH**[8–10]**.

Sensitivity to heat, with higher temperatures promoting faster viral inactivation, has been documented for SARS-CoV-2-related coronaviruses SARS-CoV **[11]** and Middle East respiratory syndrome coronavirus (MERS-CoV) **[12]**. Temperature sensitive mutants of coronavirus with heat induced loss of infectivity due to S glycoprotein processing defects were also reported **[13]**. Furthermore, inactivation of coronaviruses by deep ultraviolet (UVC) light from an artificial source is well documented **[14–17]**. Although the germicidal UVC sunlight almost never reaches the Earth, some studies reported possible inactivation of bacteria and viruses by solar radiation **[18,19]**. Given the differences in atmospheric air temperature and intensity of sunlight in different parts of the world, we hypothesize that the global spread of COVID-19 might have followed a ‘tropism’ towards countries with colder climate and less sunlight. This study, therefore, assessed whether the geographic distribution of this disease was correlated with atmospheric air temperature in different countries. Additional weather, geographical and population variables were also evaluated. Since it has been previously proposed that universal bacillus Calmette-Guérin (BCG) vaccination may be a factor influencing the outcome of COVID-19 **[20]**, we also assessed whether COVID-19 mortality rates in different countries correlate with the extent of BCG vaccination.

## 2. Materials and Methods

### 2.1. Data sources

The numbers of confirmed COVID-19 cases reported in each country between 1^st^ Jan and 31^st^ Mar 2020 were obtained from the World Health Organization’s (WHO) 71^st^ daily situational report **[2]**. The WHO defined a confirmed case of COVID-19 as “a person with laboratory confirmation of COVID-19 infection, irrespective of clinical signs and symptoms” **[21]**.

Climate data for all respective countries were obtained from the Weatherbase **[22–24]**, which provides country-by-country monthly average values for temperature and precipitation based on many years of data from the National Climatic Data Centre. The average temperature and precipitation in January, February and March were used for this study. The monthly average relative humidity was not captured in the Weatherbase dataset and hence not available for this analysis. Data on the latitude and longitude of each country’s capital city was obtained from the CSGNetwork **[25]**.

United Nations (UN) estimates of a country’s total population and median age for the year 2020 were used **[26]**. The population density of countries was obtained from https://data.worldbank.org/, and data on the national BCG policy and World Bank income group classification of individual countries were obtained from the BCG world atlas http://www.bcgatlas.org/) **[27]**. For the purpose of correlation analysis, countries were grouped into low-income, low/middle-income, high/middle-income and high-income categories.

The total number of COVID-19 cases and deaths, population (size, density, median age), climate, geographic coordinates of capital cities, income group, and BCG vaccination policies for each country are shown in Supplementary File (Table S1).

### 2.2. Study outcomes

The primary study outcome was the correlation between the total number of COVID-19 cases and atmospheric air temperature and latitude.

### 2.3. Study definitions and measures

Weather was defined as a short-term variation (minutes to weeks) of the atmosphere, and climate was defined as the weather of a place averaged over a period of time (often 30 years) **[28]**. We used monthly average climatic conditions of countries calculated over many years, as it gives a good representation of the climate in each country **[29]**. Air temperature represented the amount of heat in the air in degrees centigrade (°C) measured by a thermometer freely exposed to the atmosphere. Latitude was defined as an angular distance that defines the north– south position of the capital city of countries along the meridian and expressed as the number of degrees (°) from the equator **[30,31]**. UV levels and air temperature are higher with decreasing latitude towards the equator **[32,33]**. Precipitation was defined as any liquid or solid phase aqueous particles that originate in the atmosphere and fall down to the surface of the Earth **[34]**. Precipitation quantities were expressed in millimetres (mm). Population density refers to the number of people per square kilometre of land area. Age of BCG vaccination^3^

Submitted for publication in *Geophysical Research Letters* policy for each country was defined as the number of years between the start of a universal BCG vaccination policy up to the current year (2020). Further correlations were performed for countries with ongoing and ceased BCG vaccination programs.

### 2.4. Statistical Analysis

Spearman correlation coefficient (ρ) was obtained from bivariate correlation analysis using GraphPad Prism version 8.0.2 for Windows (GraphPad Software, San Diego, Cal., USA). Partial correlation analysis and the Kaiser-Meyer-Olkin measure of sampling adequacy (KMO) and Bartlett’s test of sphericity was performed using SPSS Statistics for Windows, version 23.0 (SPSS Inc., Chicago, Ill., USA). Missing values were excluded list-wise to create the correlation matrix in SPSS. Probability values less than 0.05 (two-sided) at 95% confidence interval were considered significant.

## 3. Results

Our analysis included a total of 748,555 confirmed COVID-19 cases worldwide that were recorded by the WHO between 1^s^t Jan and 31^st^ Mar 2020. During this period, the average air temperature in the affected countries ranged from –15.4°C in Mongolia to 31.2°C in Panama, with a world average of 15.2°C. Precipitation over this period ranged from 0.3 mm in Gambia to 316.8 mm in Fiji, with a global average of 70 mm (Supplementary File (Table S1)).

A correlation matrix from the bivariate correlation analysis showing the association of all the assessed variables with COVID-19 cases and deaths is shown in Table 1. Since most of the variables were interrelated, the multivariate model (partial correlation analysis) was not fit to obtain statistically significant associations. Atmospheric air temperature and latitude had the strongest correlation with each other (ρ = –0.88, P <0.01) (Table1). As shown in Table 1, air temperature decreased with an increase in latitude away from the equator, while the median age and income of countries increased. Another key finding here is that with an increasing latitude the number of countries with BCG vaccination at any time (past or current) decreased, whereas the age of BCG vaccination policy increased (Table 1).

**Table 1:**
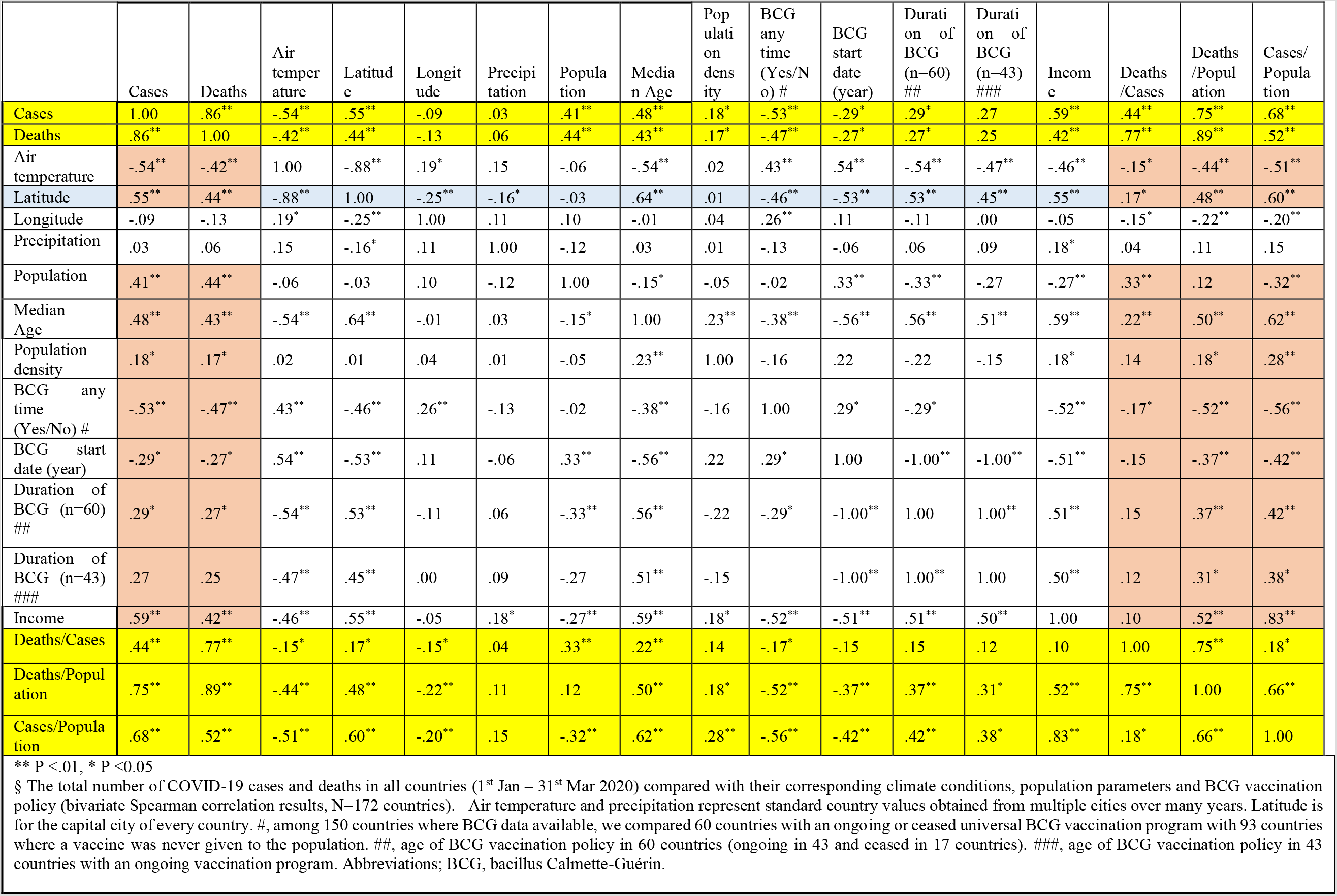
Correlation among variables^§^.

There was negative correlation between the number of COVID-19 cases and the atmospheric air temperature (Figures 1A and B). As the atmospheric temperature increased, there was a significant decrease in the number of COVID-19 cases (ρ = –0.54, 95%CI: [−0.64, –0.42]; P <0.001). In line with this observation, increasing latitude (i.e. moving from the equator towards the poles, with an implied decrease in air temperature) was associated with a significant increase in COVID-19 cases (ρ = 0.60, 95%CI: [0.48, 0.70]; P <0.001). For example, an increase in latitude from 25° (South Africa) to 50° in the United Kingdom (countries with similar COVID-19 testing strategy) corresponded to a 16-fold increase in the number of detected cases (from 2 to 32 per 100,000 population) (Supplementary File (Table S1). Atmospheric precipitation, however, did not show any correlation with the total number of COVID-19 cases (ρ = 0.02, P = 0.73) (Figure 1C).

**Figure 1:**
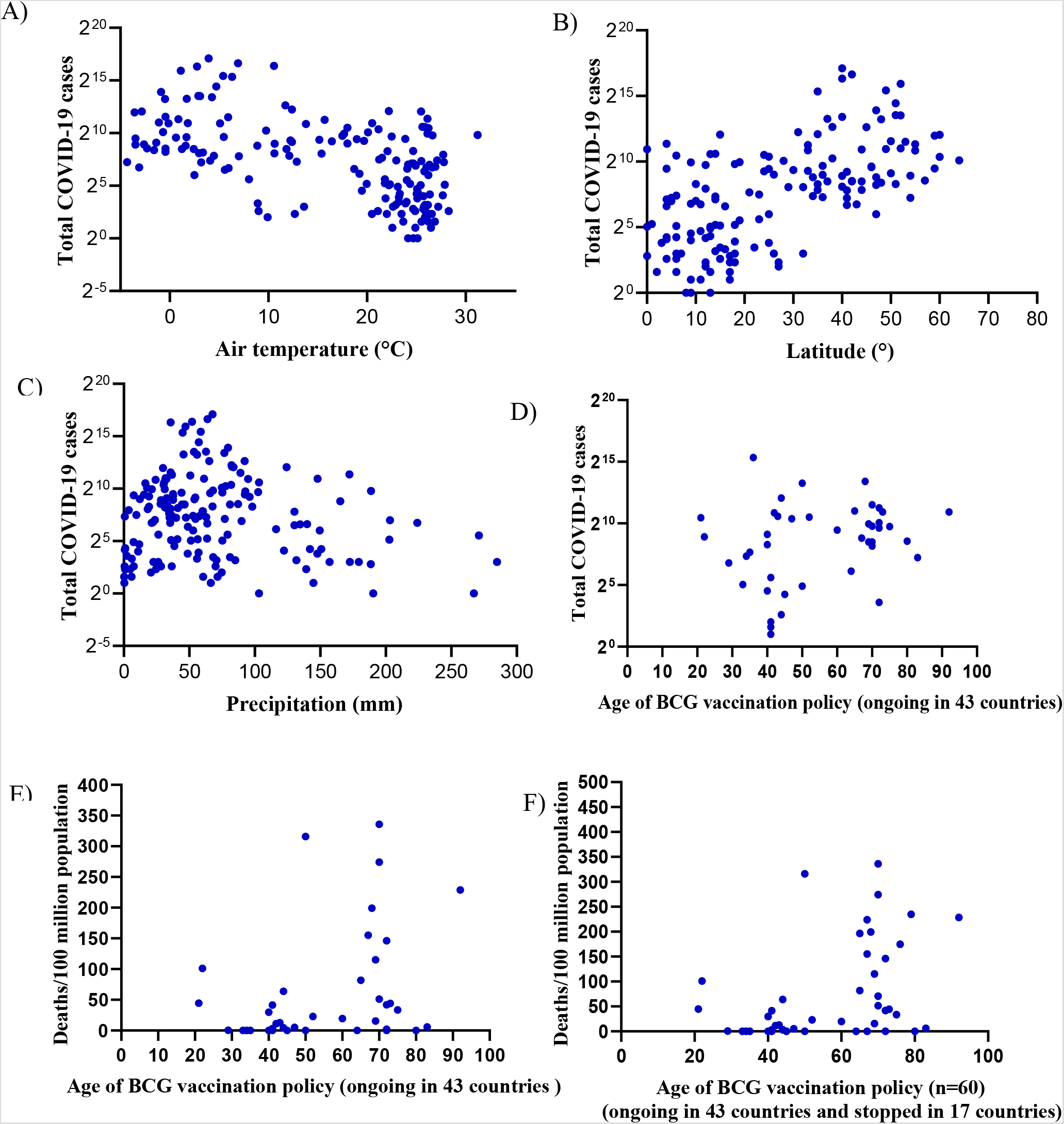
The correlation of COVID-19 with climate conditions and BCG vaccination. The total number of COVID-19 cases in different countries, and its correlation with: (A) atmospheric air temperature (ρ = –0.54, 95%CI: [−0.64, –0.42]; P<0.001); (B) latitude (ρ = 0.60, 95%CI: [0.48, 0.70]; P<0.001); (C) atmospheric precipitation (ρ = 0.03, 95%CI: [−0.13, 0.18]; P=0.73); and (D) age of BCG vaccination policy (ρ = 0.27, 95%CI: [−0.04, 0.53]; P=0.08). The correlation of COVID-19 death rate and years since the introduction of BCG vaccination(E) in 43 countries with an ongoing vaccination program (ρ = 0.31, 95%CI: [−0.003, 0.56]; P=0.05); and (F) in 60 countries (ongoing in 43 and stopped in 17 countries) (ρ = 0.37, 95%CI: [0.12, 0.57]; P=0.003). Abbreviations; °C, degree centigrade; mm, millimetre; COVID-19; coronavirus disease-19.

Universal BCG vaccination policy (past or currently ongoing) appears to be associated with decreased COVID-19 cases and deaths, and cases and deaths per population (Table 1). However, most of the countries with such vaccination policies are in the tropics (near the equator) with a high atmospheric air temperature (Table 1, Supplementary File (Table S1). However, as the age of vaccination policy (the duration in years since countries introduced BCG vaccination) increased, there was a statistically significant increase in COVID-19 cases and deaths, and cases and deaths per population (Table 1, Figure 1E and F). Nevertheless, the^4^ Submitted for publication in *Geophysical Research Letters* duration of BCG vaccination did not decrease COVID-19 deaths among cases (ρ = 0.15, 95%CI: [-0.12, 0.39]; P = 0.27) (Table 1).

## 4. Discussion

This study has demonstrated the association between the prevalence of COVID-19 and air temperature. An increasing number of COVID-19 cases were seen at geographic locations with lower atmospheric air temperature and/or increasing latitude. Although we could not confirm causation, the atmospheric air temperature was in some way related to the global distribution of COVID-19. From a careful interpretation of Table 2 and existing scientific evidence, we also show that the confounding factors (median age of population, income level, and BCG vaccination) were the least likely causes for the tropism of COVID-19 disease towards the poles of the Earth.

There are two possible explanations as to why temperature may limit transmission. First, high air temperature in the tropics may directly inactivate SAR-CoV-2 on surfaces, the ground and in aerosols, decreasing rate of transmission. Indeed, the transmitting ability (effective reproductive number) of SARS CoV-2-infected persons was previously shown to decrease with increasing air temperature **[35]**. This explanation is also consistent with the results of an earlier study that reported a local decrease in the incidence of COVID-19 in China with increasing air temperature**[36]**. SARS CoV-2 can remain stable on surfaces, for example, for up to 72 hours **[37]**; lower environmental temperature may be an important factor allowing that.

Secondly, decrease in COVID-19 incidence with decreasing latitude (and the corresponding increase in temperature) could be due to higher UV radiation from the sun – the closer to the equator, the higher the UV radiation levels **[38]**. High sensitivity of coronaviruses to UV light was documented in a study that found a greater sensitivity of a SAR-CoV-1 surrogate **[12]** called murine hepatitis virus (MHV) to UV light than adenovirus **[39]**. Furthermore, UVA, in combination with the light-activated DNA and RNA crosslinking agent amotosalen, was shown to effectively inactivate MERS-CoV infectious particles**[16]**. The other possibility is that vitamin-D deficiency and an increased susceptibility to infection could be the responsible factor in areas away from sunlight**[40]**.

The confounding variables (level of income, BCG vaccination, and median age of population) also showed a correlation with an increased incidence of COVID-19. However, this was likely because countries with good income, higher median age and lower incidence of tuberculosis (hence no BCG vaccination) are generally located at higher latitude (and therefore lower air temperature) (Table 1, Supplementary File (Table S1)). The results of our analysis appear to contrast with the findings of a previous study that reported a correlation of universal BCG vaccination with a decreased number of COVID-19 deaths per country **[20]**. In contrast, our study (using a larger sample) showed increased COVID-19 mortality in countries with a longer duration in years since mass BCG vaccination was introduced. Again, the likely explanation for this is that countries with older BCG vaccination policies tend be located at higher latitude, i.e. further away from the equator (Table 1). It is important to note that the positive correlation between COVID-19 deaths and the age of the BCG vaccination policy reached a statistical significance when the 17 countries that stopped BCG vaccination were added (14 were European countries that had low temperature and greater age of BCG vaccination policy) (Figures 1E and F, Supplementary File (Table S1)). Altogether, this suggests that there is no link between BCG vaccination and COVID-19 mortality. In fact, the 3 of 13 duration of BCG vaccination did not correlate with the fatality rate (Deaths/cases) of COVID-19 (Table 1).

Higher median age of population in countries closer to the Earth’s poles could be another plausible factor linked to an increased number of COVID-19 cases in these locations. However, as shown in Table 1, the median age association with COVID-19 cases per capita was stronger (ρ=0.62, P<0.01) than its association with COVID deaths per total cases (ρ=0.22, P<0.01). This does not align with the extensive available evidence that increasing age of patients has a much stronger association with the case fatality rate than with the number of cases; indeed, preferential infection of older people was not reported, and in the WHO’s 7^th^ daily situational report, the median age among all COVID-19 infected people was 45 years. We can therefore rule out age as an influencing factor in this analysis. Similarly, as discussed above, income was correlated with COVID-19 cases due to its association with latitude (countries with higher average income were further away from the equator) (Table 1). Hence, the variables other than temperature and latitude were most likely confounders. Population density, however, has a clear correlation with an increased COVID-19 cases per population as it was not correlated with latitude (Table 1).

This study has several limitations. First, since most of the variables were already interrelated, the multivariate model (partial correlation analysis) did not give statistically significant associations. The second major limitation is that COVID-19 epidemic is still unfolding in most of the tropical countries particularly Africa. Nevertheless, considering case rate increase, case incidence rate was lower in some African countries relative to European countries using the same testing strategy. There could also be many other unidentified factors behind the case incidence rate reported in each country.

## 5. Conclusions

The geographic spread of COVID-19 is negatively correlated with atmospheric air temperature and/or sunlight. Lower atmospheric air temperature may have played an important role in the global spread of COVID-19. A longer duration of mass BCG vaccination did not decrease COVID-19 fatality rate. Our analysis justifies further studies of the effect of the air temperature on the survival and infectivity of SAR-CoV-2.

## Data Availability

All the data we used to make this paper can be accessed online in the following webpages. Data on COVID-19 cases and deaths of all countries can be downloaded here (https://www.who.int/emergencies/diseases/novel-coronavirus-2019/situation-reports). Average climate records of countries are found at https://www.weatherbase.com/ whereas data on the latitude of capital cities can be accessed here (http://www.csgnetwork.com/llinfotable.html). Data on the total population and median age of countries was taken from https://www.un.org/en/development/desa/population/index.asp and https://www.un.org/en/development/desa/population/index.asp respectively.

## Supplementary Materials

The following are available online at, Table S1: The total number of COVID-19 cases and deaths, population (size, density, median age), climate, geographic coordinates of capital cities, income group, and BCG vaccination policies of countries (=172 countries).

## Author Contributions

Conceptualization, Y.B. and W.B.; methodology, Y.B. and W.B.; validation, Y.B., A.R., W.B, E.A., W.S., A.M. and A.B.; formal analysis, Y.B.; investigation, Y.B., W.B, A.M. and A.B.; data curation, A.M., Y.B., and W.B.; writing—original draft preparation, Y.B.; writing—review and editing, Y.B., A.R., W.B, E.A., W.S., A.M. and A.B.; visualization, Y.B., A.R., W.B, E.A., W.S., A.M. and A.B.; supervision, W.B.; project administration, W.B. and Y.B. All authors have read and agreed to the published version of the manuscript.

## Funding

This research received no external funding.

## Acknowledgments

We thank Sibhat Mequanint, Yeshambaw Belachew, Yohannes Gishen and Getu Mequanint for their contribution in the data collection.

## Conflicts of Interest

The authors declare no conflict of interest.

## Plain Language Summary

Severe acute respiratory syndrome coronavirus 2 (SARS-CoV-2) is an enveloped virus that could be sensitive to certain adverse factors in the external environment. For example, the virus needs to survive and remain infectious on various surfaces for effective human to human transmission. We assessed whether the global country-to-country variation of coronavirus disease-19 incidence and mortality could correlate with atmospheric air temperature, precipitation, and additional geographic and population variables. We found that the geographic spread of COVID-19 is negatively correlated with atmospheric air temperature and/or sunlight. Lower atmospheric air temperature may have played an important role in the global spread of COVID-19.

## Notes

### Competing Interest Statement

The authors have declared no competing interest.

### Author Declarations

This is an ecological study and we used secondary data that is freely available on the internet. Individual subjects cannot be identified in any way. Hence, this manuscript fulfills the exemption criteria for IRB oversight.

